# Changing trends in proportional incidence and net survival of screened and nonscreened breast cancers among women in England

**DOI:** 10.1101/19003202

**Authors:** Haiyan Wu, Kwok Wong, Shou-En Lu, John Broggio, Lanjing Zhang

## Abstract

**Background:** Uptake of breast cancer screening has been decreasing in England since 2007, and may increase proportional incidence of nonscreened cancers. However, recent trends in proportional incidence and net-survivals of screened and nonscreened breast cancers are unclear.

**Methods:** We extracted population-based proportional incidence and age-standardized 5-year net-survivals from Public Health England, for English women with invasive breast cancer diagnosed during 1995-2011 (linked to death certificates, followed through 2016). Piecewise log-linear models with change-point/joinpoint were used to estimate temporal trends. We conducted a quasi-experimental study to test the hypothesis that the trend-change year of proportional incidence coincided with that of 5-year net-survival.

**Results:** Among 254,063 women in England with invasive breast cancer diagnosed during 1995-2011, there was downward-to-upward trend-change in proportional incidence of nonscreened breast cancers (annual percent change[APC]=5.6 after 2007 versus APC=-3.5 before 2007, *P*<0.001) in diagnosis-year 2007, when steeper upward-trend in age-standardized 5-year net survival started (APC=5.7 after 2007/2008 versus APC=0.3 before 2007/2008, *P*<0.001). Net-survival difference of screened versus nonscreened cancers also significantly narrowed (18% in 2007/2008 versus 5% in 2011). Similar associations were found in all strata of race, cancer stage, grade and histology, except in Black patients or patients with stage I, stage III, or grade I cancer.

**Conclusions:** The downward-to-upward trend-change in proportional incidence of nonscreened breast cancers is associated with steeper upward-trend in age-standardized 5-year net survival among English women in recent years. Survival benefits of breast cancer screening appear decreasing in recent years. The data support reduction of breast cancer screening in some patients.

## Introduction

Cancer screening is adopted worldwide for breast cancer prevention and control.^1^ The uptake of breast cancer screening has been decreasing among women in England since 2007, from 73.2% among women aged 50-70 years in 2007-2008 to 70.5% in 2017-2018.^2,3^ Given widely accepted benefits of cancer screening,^4,5^ the decrease in screening may increase the proportional incidence of nonscreened breast cancers and suppress improvement of their survivals. However, the long-term trends in proportional incidence of screened and nonscreened breast cancers are largely unknown, despite an overall upward trend of breast cancer incidence in England.^6,7^

The benefits and harms of breast cancer screening are controversial, although benefits appear to outweigh harms. ^5,8-11^ Thus, the recent decrease in uptakes of breast cancer screening in England may also link to different changes in breast cancer survival of nonscreened and screened patients. However, the trends in net survival of screened and nonscreened invasive breast cancers are unclear among women in England, while the overall net survivals have been increased in recent years.^6,7^ Therefore, using data from Public Health England, we estimated these 5-year net survival trends during 1995-2016. We also conducted a quasi-experimental study to examine whether the trend change in proportional incidence of nonscreened invasive breast cancers is associated with trend change in age-standardized 5-year net survival of these cancers among women in England. Subgroup analyses by cancer stage, histology, cancer grade and patient race were also performed. This quasi-experimental study may help better understand the benefits of breast cancer screening in recent years.

## Methods

We requested the aggregated data of proportional incidence and age-standardized 5-year net survival of invasive breast cancer by various factors, which were prepared for, calculated using Stata (version 15, StataCorp LLC, TX), and released by the National Cancer Registration and Analysis Service within Public Health England (PHE).^6,12^ The database has been used for studying breast, pediatric and colorectal cancers.^13-15^ The invasive breast cancer was classified according to the International Statistical Classification of Diseases 10th Revision (ICD-10) and by morphology and behavior codes in the International Classification of Diseases for Oncology, Second Edition (ICD-O-2). The net survival is a ratio calculated by dividing the overall/observed survival of cancer patients over that of the general population using the Pohar-Perme estimator.^16^ The net survival used here adjusts the survival of breast cancer patients with that of the general population using an updated, smoothed life table.^17^ Age-standardization was performed using the International Cancer Survival Standard age-weightings.^18^ We included all qualified invasive breast cancers (site and morphology. Primary site-labeled: breast) in England diagnosed during 1995-2011 (released in February 2019). The exclusion criteria included: Death certificate only, autopsy only, or alive with no survival time; Exclusion to match the expected survival table: age value not found in table, invalid year and values not found for other variables. Since we used this existing, de-identified, publicly available dataset, no Institutional Review Board (IRB) review was required.

We classified the cancer stage using a TNM-based staging system defined by Cancer Research UK.^19^ The cancer histology was classified and categorized using the International-Classification of Disease for Oncology (ICD-O)-2,^20^ according to the pathology diagnosis in medical charts. We grouped the tumors into invasive ductal carcinoma (IDC, ICD-O-2 8500/3), invasive lobular carcinoma (ILC, ICD-O-2 8520/3), mixed invasive ductal and lobular carcinoma (MDLC, ICD-O-2 8522/3) and non-ductal non-lobular carcinomas (all other codes) for the primary analyses. We stratified the proportional incidence and age-standardized 5-year net-survivals by diagnosis year, race, histology, cancer stage and cancer grade among women with screened or nonscreened breast cancer. We calculated proportional incidence using stratum’s incident case number divided by the number of all strata’s incident cases.

In the quasi-experimental study, we identified and compared the changing points of the trends in proportional incidence and age-standardized 5-year net survival, respectively, using piece-wise log-linear models of the Joinpoint program (Version 4.6.0.0., Statistical Research and Applications Branch, National Cancer Institute, Bethesda, MD).^21^ We employed the following setups for analyses: standard errors (provided) option for Heteroscedastic Errors Option (Weighted Least Squares); grid search method with 2 as the minimal number of observations from a joinpoint to either end of the data (excluding the first or last joinpoint if it falls on an observation) and the minimal number of observations between two joinpoints (excluding any joinpoint if it falls on an observation).^21,22^ The model selection for the best-fit joinpoint was based on permutation tests with overall significance level at 0.05. We also compared the trends/slopes among the strata using the pairwise comparison function of Joinpoint program.^22^ On very rare occasions (<1%), age-standardized net-survivals were unavailable due to missing data, and those data points would be omitted in the analysis. All *P* values were 2-sided, and would be considered statistically significant when <0.05.

## Results

### Trends in the proportional incidence of invasive breast cancer among women in England diagnosed during 1995-2011

Among the 254,063 women in England with invasive breast cancer diagnosed during 1995-2011 (183,018 [72.0%] IDC, 30,323 [11.9%] ILC, 9,324 [3.7%] MDLC, and 31,398 [12.4%] others), 122,870 (48.8%) were screened cancers overall (**Table 1**). The proportional incidence of screened breast cancer (versus nonscreened) was significantly different by diagnosis year, race, histology, stage and tumor grade (**Table 1**).We found a joinpoint in the proportional incidence of nonscreened breast cancer in 2007, when the proportion of screened cancer increased from 37.3% to 57.0% during 1995-2007 (APC[95% Confidence interval, CI]=4.0 [3.1 to 5.0], *P*<0.001) but decreased from 57.0% to 47.6% afterward (APC [95% CI]=-5.3 [-8.2 to -2.3], *P*=0.002, **Table 2**). The same joinpoint of 2007 was also identified in the trend of proportional incidence of nonscreened breast cancer (**Figure**). The trends in proportional incidence of screened breast cancer too differed by histology, stage and cancer grade, but not race (**Table 2**).

**Table 1.** Characteristics of the screened and nonscreened breast cancers among the Women in England, 1995-2016

|  | Nonscreened |  | Screened |  | All |  | *P* |
| --- | --- | --- | --- | --- | --- | --- | --- |
|  | N (%) | NS, % | N (%) | NS, % | N | NS, % |  |
| **Year** |  |  |  |  |  |  | <0.001 |
| 1995 | 4665 (62.7) | 77 | 2771 (37.3) | 96 | 7436 | 84 |  |
| 1996 | 5175 (62.4) | 77 | 3122 (37.6) | 93 | 8297 | 83 |  |
| 1997 | 7036 (65.3) | 77 | 3744 (34.7) | 98 | 10780 | 84 |  |
| 1998 | 6484 (61.4) | 78 | 4071 (38.6) | 100 | 10555 | 86 |  |
| 1999 | 6156 (57.4) | 79 | 4563 (42.6) | 99 | 10719 | 88 |  |
| 2000 | 5794 (53.8) | 78 | 4966 (46.2) | 101 | 10760 | 88 |  |
| 2001 | 5781 (52.0) | 80 | 5331 (48.0) | 96 | 11112 | 88 |  |
| 2002 | 5894 (50.7) | 78 | 5726 (49.3) | 100 | 11620 | 89 |  |
| 2003 | 6585 (49.7) | 79 | 6671 (50.3) | 99 | 13256 | 89 |  |
| 2004 | 7132 (47.8) | 79 | 7778 (52.2) | 99 | 14910 | 89 |  |
| 2005 | 7191 (45.5) | 79 | 8599 (54.5) | 100 | 15790 | 90 |  |
| 2006 | 8597 (46.2) | 77 | 9996 (53.8) | 101 | 18593 | 89 |  |
| 2007 | 7907 (43.0) | 81 | 10479 (57.0) | 99 | 18386 | 91 |  |
| 2008 | 8965 (44.4) | 81 | 11241 (55.6) | 99 | 20206 | 91 |  |
| 2009 | 9310 (46.3) | 86 | 10795 (53.7) | 102 | 20105 | 94 |  |
| 2010 | 12779 (53.3) | 88 | 11202 (46.7) | 101 | 23981 | 94 |  |
| 2011 | 12991 (52.4) | 95 | 11815 (47.6) | 100 | 24806 | 97 |  |
| **Race** |  |  |  |  |  |  | <0.001 |
| Black | 973 (64.6) | 81 | 533 (35.4) | 89 | 1506 | 85 |  |
| Other | 54387 (50.9) | 76 | 52543 (49.1) | 97 | 106930 | 86 |  |
| White | 73068 (51.2) | 87 | 69727 (48.8) | 101 | 142795 | 93 |  |
| **Histology** |  |  |  |  |  |  | <0.001 |
| IDC | 91671 (50.1) | 84 | 91347 (49.9) | 99 | 183018 | 91 |  |
| ILC | 15960 (52.6) | 86 | 14363 (47.4) | 98 | 30323 | 92 |  |
| MDLC | 3867 (41.5) | 86 | 5457 (58.5) | 97 | 9324 | 92 |  |
| Other | 18540 (59.1) | 70 | 12858 (41.0) | 97 | 31398 | 80 |  |
| **Tumor Stage\*** |  |  |  |  |  |  | <0.001 |
| Stage1 | 25857 (36.2) | 97 | 45636 (63.8) | 102 | 71493 | 99 |  |
| Stage2 | 29271 (61.7) | 87 | 18162 (38.3) | 95 | 47433 | 90 |  |
| Stage3 | 6119 (75.3) | 68 | 2009 (24.7) | 88 | 8128 | 73 |  |
| Stage4 | 4193 (85.0) | 31 | 743 (15.1) | 72 | 4936 | 38 |  |
| **Tumor grade\*** |  |  |  |  |  |  | <0.001 |
| Grade1 | 16078 (32.4) | 96 | 33603 (67.6) | 103 | 49681 | 99 |  |
| Grade2 | 52877 (47.1) | 90 | 59422 (52.9) | 100 | 112299 | 95 |  |
| Grade3 | 44455 (64.9) | 75 | 24069 (35.1) | 92 | 68524 | 81 |  |
| **Total** |  |  |  |  |  |  |  |
|  | 13038 (51.2) | 82 | 122870 (48.8) | 99 | 254062 | 90 |  |

NS, age-standardized 5-year net survival; IDC, invasive ductal carcinoma; ILC, invasive lobular carcinoma; MDLC, mixed ductal and lobular carcinoma; *P*, chi-square test for the within group differences; \* data were missing in some cases.

**Table 2.** Trends in proportion of screened and nonscreened breast cancers diagnosed during 1995-2011 among the women in England

|  | Screened, % |  |  |  | Nonscreened, % |  |  |  |
| --- | --- | --- | --- | --- | --- | --- | --- | --- |
|  | 1995 | 2011 | APC (95% CI) | P | 1995 | 2011 | APC (95% CI) | P |
| **All** | 37.3 | 57.0* | 4.0 (3.1 to 5.0) |  | 62.7 | 43.0* | -3.5 (-4.2 to -2.8) |  |
|  | 57.0* | 47.6 | -5.3 (-8.2 to -2.3) |  | 43.0* | 52.4 | 5.6 (2.2 to 9.1) |  |
| **Race** | 1998 |  |  |  |  |  |  |  |
| Black | 27.3 | 37.1 | 2.3 (-0.4 to 5.1) | reference | 72.7 | 62.9 | -1.8 (-3.3 to -0.3) | reference |
| Other | 36.9 | 60.0# | 3.4 (0.5 to 6.3) | 0.985 | 63.1 | 40.0# | -4.1 (-6.6 to -1.5) | 0.879 |
|  |  |  | -10.0 (-21.9 to |  |  |  | 12.9 (-4.2 to |  |
|  | 60.0# | 46.8 | 3.6) |  | 40.0# | 53.2 | 33.0) |  |
| White | 41.0 | 52.9^ | 4.7 (1.9 to 7.5) | 0.963 | 59.0 | 45.8* | -3.5 (-5.5 to -1.4) | 0.783 |
|  | 52.9^ | 48.2 | -3.0 (-5.9 to -0.1) |  | 45.8* | 51.8 | 4.2 (-0.8 to 9.5) |  |
| **Histology** |  |  |  |  |  |  |  |  |
| IDC | 39.8 | 57.4* | 3.6 (2.6 to 4.5) | 0.736 | 60.2 | 42.7* | -3.3 (-4.1 to -2.5) | 0.998 |
|  | 57.4* | 48.5 | -4.8 (-7.7 to -1.8) |  | 42.7* | 51.6 | 5.3 (1.7 to 9.0) |  |
| ILC | 36.9 | 56.8* | 4.2 (3.5 to 4.9) | reference | 63.1 | 43.2* | -3.3 (-3.9 to -2.8) | reference |
|  | 56.8* | 45.5 | -5.6 (-7.7 to -3.5) |  | 43.2* | 54.5 | 5.4 (2.9 to 8.0) |  |
| MDLC | 69.5 | 62.7* | 1.9 (0.3 to 3.5) | 0.014 | 30.5 | 35.1* | -3.6 (-5.9 to -1.3) | 0.176 |
|  | 62.7* | 52.4 | -8.4 (-17.3 to 1.5) |  | 35.1* | 47.6 | 9.6 (0.9 to 18.9) |  |
| Other | 27.8 | 50.4& | 7.7 (5.9 to 9.4) | <0.001 | 72.2 | 51.0* | -4.1 (-4.9 to -3.3) | 0.005 |
|  | 50.4& | 42.1 | -2.8 (-4.3 to -1.1) |  | 51.0* | 57.9 | 2.9 (1.0 to 4.8) |  |
| **Tumor stage** |  |  |  |  |  |  |  |  |
| Stage1 | 49.1 | 27.1# | -5.2 (-6.6 to -3.7) | reference | 50.9 | 71.9* | 3.8 (2.8 to 4.9) | reference |
|  | 27.1# | 31.4 | 5.6 (-4.9 to 17.3) |  | 71.9* | 68.6 | -1.7 (-4.1 to 0.7) |  |
| Stage2 | 73.3 | 54.4* | -3.1 (-4.1 to -2.1) | 0.024 | 26.7 | 45.7^ | 5.9 (3.5 to 8.4) | <0.001 |
|  | 54.4* | 59.5 | 4.1 (-0.4 to 8.8) |  | 45.7^ | 40.6 | -3.7 (-7.5 to 0.3) |  |
| Stage3 | 88.6 | 70.1 | -1.8 (-2.5 to -1.1) | 0.003 | 11.4 | 32.7^ | 14.5 (7.6 to 21.9) | <0.001 |
|  |  |  |  |  | 32.7^ | 29.9 | -2.5 (-9.0 to 4.4) |  |
| Stage4 | 90.1 | 77.9 | -1.0 (-1.3 to -0.7) | 0.048 | 9.9 | 22.1 | 6.5 (4.7 to 8.4) | <0.001 |
| **Tumor Grade** |  |  |  |  |  |  |  |  |
| Grade1 | 58.0 | 71.5* | 2.7 (2.1 to 3.3) | reference | 42.0 | 24.9* | -5.1 (-6.2 to -3.9) | reference |
|  | 71.5* | 68.4 | -3.1 (-5.2 to -0.9) |  | 24.9* | 31.6 | 7.0 (1.1 to 13.2) |  |
| Grade2 | 40.2 | 61.6* | 4.3 (3.4 to 5.2) | 0.002 | 59.8 | 38.4* | -4.3 (-5.1 to -3.5) | 0.018 |
|  | 61.6* | 51.3 | -5.2 (-7.6 to -2.7) |  | 38.4* | 48.7 | 6.4 (2.9 to 10.1) |  |
| Grade3 | 26.1 | 41.5# | 4.1 (3.0 to 5.1) | 0.001 | 73.9 | 58.5# | -2.1 (-2.6 to -1.6) | 0.001 |
|  | 41.5# | 33.3 | -9.4 (-14.7 to -3.8) |  | 58.5# | 66.7 | 5.7 (1.9 to 9.5) |  |

IDC, invasive ductal carcinoma; ILC, invasive lobular carcinoma; MDLC, mixed ductal and lobular carcinoma; P, P-values of parallelism test for the within group differences (<0.05 indicates different slopes/trends compared with the reference); APC, annual percentage change; CI, confidence intervals; symbols indicate the diagnosis year corresponding to a joinpoint when two slopes/trends intercept, including & for 2004, ^ for 2006, \* for 2007 and # for 2008.

**Figure.**
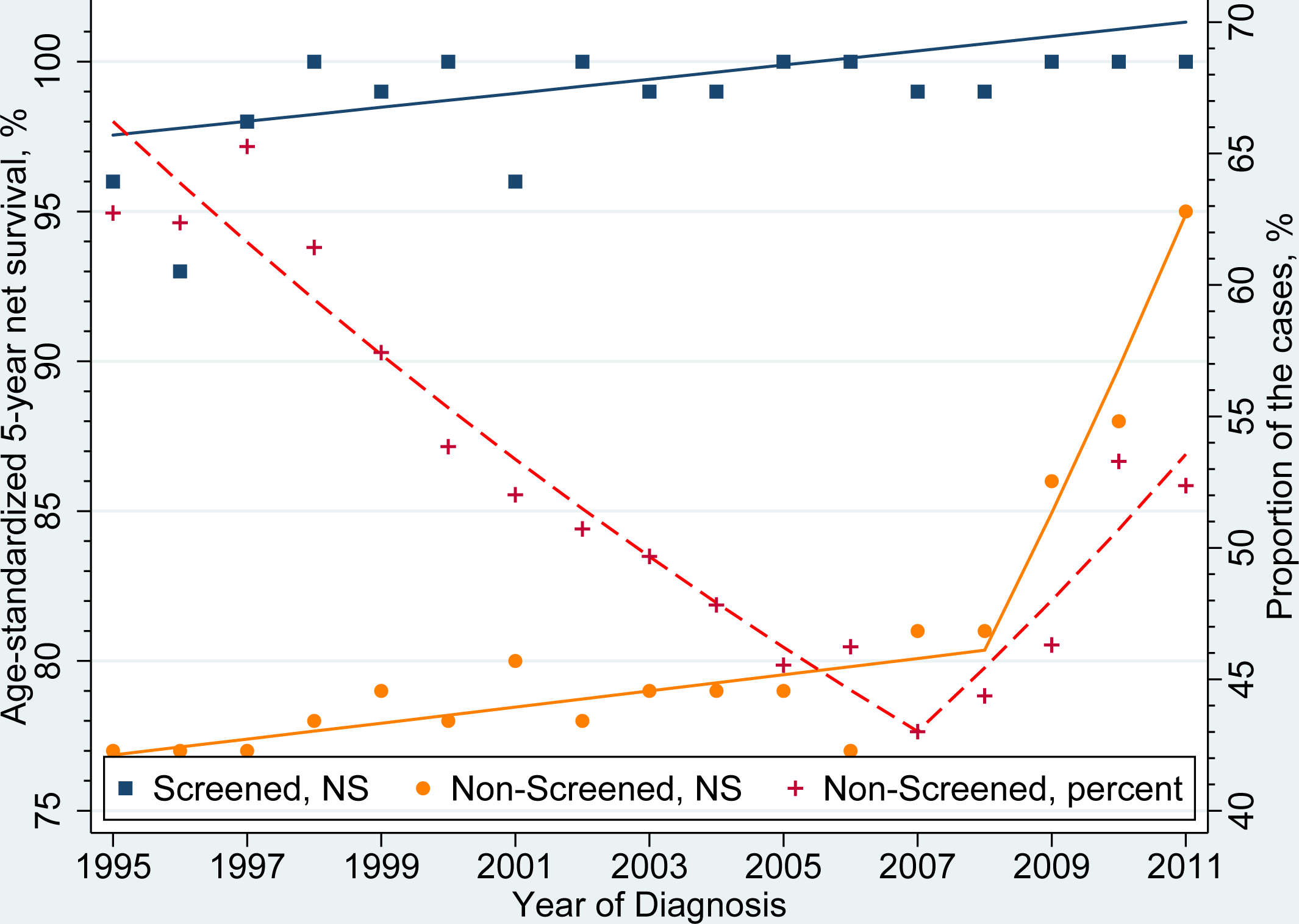
Trends in proportion and age-standardized 5-year net survival of screened and nonscreened breast cancer diagnosed during 1995-2011 among women in England (followed through 2016). The dots show individual data points, while the lines show piece-wise log-linear trends of the best-fit model which were identified using the Joinpoint program. There was a downward trend in the proportion of incident nonscreened breast cancers (Annual percentage change [APC], 95% confidence intervals [CI]= -3.5 (−4.2 to -2.8), *P*<0.001, **Red color and the right Y axis**) during the diagnosis years of 1995-2007, followed by an upward trend (APC, 95% CI=5.6 (2.2 to 9.1), *P*=0.003) afterward. The screened breast cancers had an upward trend in age-standardized 5-year net-survival (APC, 95% CI= 0.4 (1.0 to 2.9), *P*=0.01, **Orange color**), while the nonscreened breast cancer had an upward trends during the diagnosis years of 1995-2008 (APC, 95% CI=0.6 [1.0 to 3.1], *P*=0.009, **Blue color**), followed by an even steeper upward trend afterward (APC, 95% CI=7.1 [1.0 to 9.2], *P*<0.001). The jointpoint of the trends in the proportion of cancer cases and that in age-standardized 5-year net-survival were similar (2007 and 2008, respectively) among the women with nonscreened invasive breast cancer.

### Trends in the age-standardized 5-year net-survival of screened and nonsreened breast cancers diagnosed among women in England during 1995-2011 (followed through 2016)

The age-standardized 5-year net survival of screened cancer was higher than that of nonscreened cancer, while the difference was significantly decreasing for the cancers diagnosed during 2007-2011 (19% difference for cancers diagnosed in 1995 versus 18% and 5% for those diagnosed in 2007/2008 and 2011, respectively, *P*_*parallelism*_<0.001, **Table 3**). There were upward trends in the age-standardized 5-year net-survival of screened and nonsreened breast cancers diagnosed among women in England, while a steeper upward trend was seen for the cancers diagnosed after 2007/2008 (2007 and 2008 had the same survivals, **Figure**). This joinpoint appeared to coincide with the joinpoint of proportional incidence of nonscreened breast cancer, and supports our hypothesis. For both screened and nonscreened breast cancers, the trends of age-standardized 5-year net-survival differed by race, histology, stage and tumor grade (**Table 3**). Compared with screened breast cancers, nonscreened breast cancers also showed different trends in the age-standardized 5-year net-survival by these factors, although some strata did not show trend difference, such as White race and stage 2 cancer (*P*_*parallelism*_=0.454 and 0.053, respectively. **Table 3**).

**Table 3.**
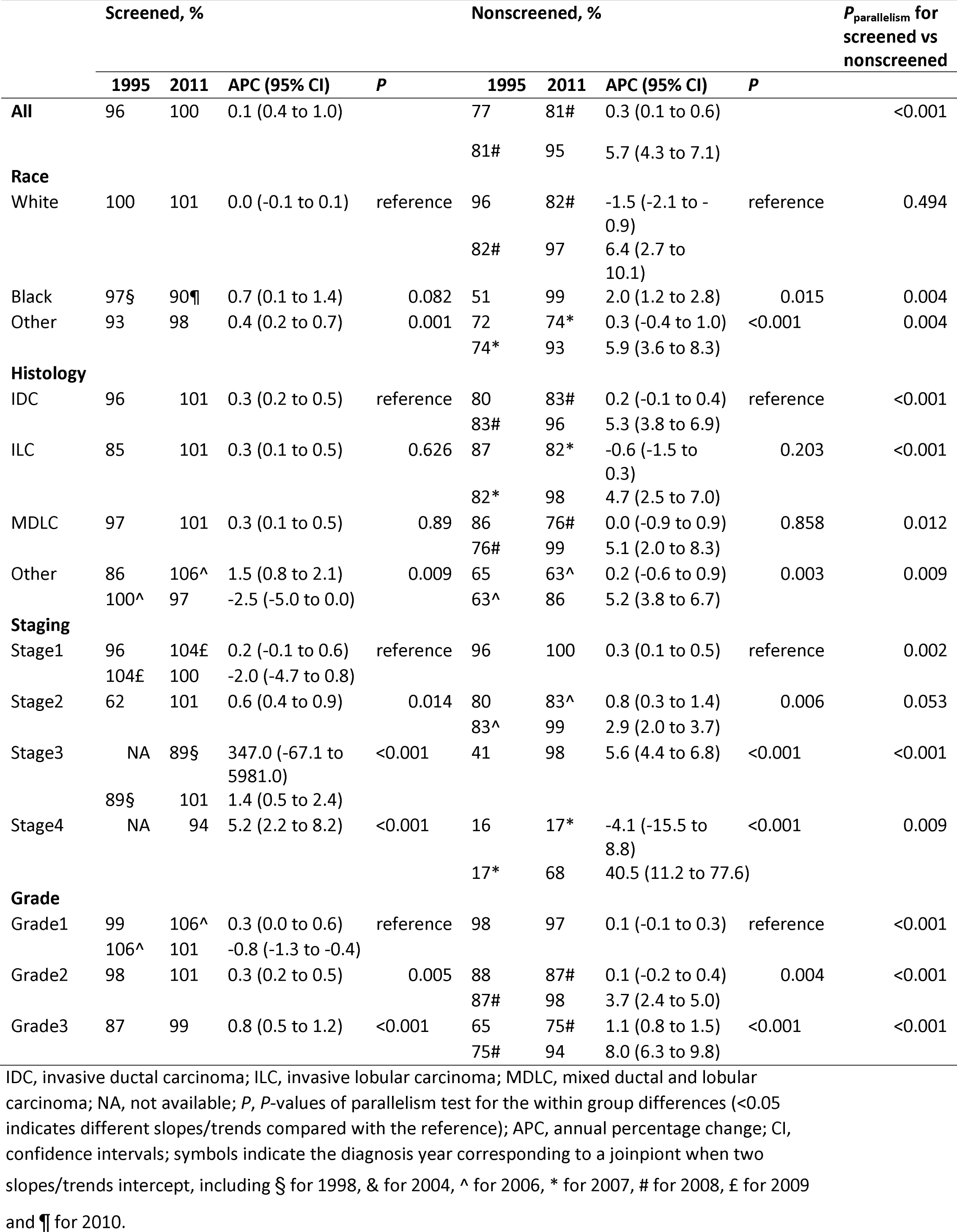
Trends in age-standardized 5-year net survival of screened and nonscreened breast cancers diagnosed during 1995-2011 among the women in England

|  | Screened, % |  |  |  | Nonscreened, % |  |  |  | *P*<sub>parallelism</sub> for screened vs nonscreened |
| --- | --- | --- | --- | --- | --- | --- | --- | --- | --- |
|  | 1995 | 2011 | APC (95% CI) | *P* | 1995 | 2011 | APC (95% CI) | *P* |  |
| **All** | 96 | 100 | 0.1 (0.4 to 1.0) |  | 77 | 81# | 0.3 (0.1 to 0.6) |  | <0.001 |
|  |  |  |  |  | 81# | 95 | 5.7 (4.3 to 7.1) |  |  |
| **Race** |  |  |  |  |  |  |  |  |  |
| White | 100 | 101 | 0.0 (-0.1 to 0.1) | reference | 96 | 82# | -1.5 (-2.1 to -0.9) | reference | 0.494 |
|  |  |  |  |  | 82# | 97 | 6.4 (2.7 to 10.1) |  |  |
| Black | 97§ | 90¶ | 0.7 (0.1 to 1.4) | 0.082 | 51 | 99 | 2.0 (1.2 to 2.8) | 0.015 | 0.004 |
| Other | 93 | 98 | 0.4 (0.2 to 0.7) | 0.001 | 72 | 74* | 0.3 (-0.4 to 1.0) | <0.001 | 0.004 |
|  |  |  |  |  | 74* | 93 | 5.9 (3.6 to 8.3) |  |  |
| **Histology** |  |  |  |  |  |  |  |  |  |
| IDC | 96 | 101 | 0.3 (0.2 to 0.5) | reference | 80 | 83# | 0.2 (-0.1 to 0.4) | reference | <0.001 |
|  |  |  |  |  | 83# | 96 | 5.3 (3.8 to 6.9) |  |  |
| ILC | 85 | 101 | 0.3 (0.1 to 0.5) | 0.626 | 87 | 82* | -0.6 (-1.5 to 0.3) | 0.203 | <0.001 |
|  |  |  |  |  | 82* | 98 | 4.7 (2.5 to 7.0) |  |  |
| MDLC | 97 | 101 | 0.3 (0.1 to 0.5) | 0.89 | 86 | 76# | 0.0 (-0.9 to 0.9) | 0.858 | 0.012 |
|  |  |  |  |  | 76# | 99 | 5.1 (2.0 to 8.3) |  |  |
| Other | 86 | 106^ | 1.5 (0.8 to 2.1) | 0.009 | 65 | 63^ | 0.2 (-0.6 to 0.9) | 0.003 | 0.009 |
|  | 100^ | 97 | -2.5 (-5.0 to 0.0) |  | 63^ | 86 | 5.2 (3.8 to 6.7) |  |  |
| **Staging** |  |  |  |  |  |  |  |  |  |
| Stage1 | 96 | 104£ | 0.2 (-0.1 to 0.6) | reference | 96 | 100 | 0.3 (0.1 to 0.5) | reference | 0.002 |
|  | 104£ | 100 | -2.0 (-4.7 to 0.8) |  |  |  |  |  |  |
| Stage2 | 62 | 101 | 0.6 (0.4 to 0.9) | 0.014 | 80 | 83^ | 0.8 (0.3 to 1.4) | 0.006 | 0.053 |
|  |  |  |  |  | 83^ | 99 | 2.9 (2.0 to 3.7) |  |  |
| Stage3 | NA | 89§ | 347.0 (-67.1 to 5981.0) | <0.001 | 41 | 98 | 5.6 (4.4 to 6.8) | <0.001 | <0.001 |
|  | 89§ | 101 | 1.4 (0.5 to 2.4) |  |  |  |  |  |  |
| Stage4 | NA | 94 | 5.2 (2.2 to 8.2) | <0.001 | 16 | 17* | -4.1 (-15.5 to 8.8) | <0.001 | 0.009 |
|  |  |  |  |  | 17* | 68 | 40.5 (11.2 to 77.6) |  |  |
| **Grade** |  |  |  |  |  |  |  |  |  |
| Grade1 | 99 | 106^ | 0.3 (0.0 to 0.6) | reference | 98 | 97 | 0.1 (-0.1 to 0.3) | reference | <0.001 |
|  | 106^ | 101 | -0.8 (-1.3 to -0.4) |  |  |  |  |  |  |
| Grade2 | 98 | 101 | 0.3 (0.2 to 0.5) | 0.005 | 88 | 87# | 0.1 (-0.2 to 0.4) | 0.004 | <0.001 |
|  |  |  |  |  | 87# | 98 | 3.7 (2.4 to 5.0) |  |  |
| Grade3 | 87 | 99 | 0.8 (0.5 to 1.2) | <0.001 | 65 | 75# | 1.1 (0.8 to 1.5) | <0.001 | <0.001 |
|  |  |  |  |  | 75# | 94 | 8.0 (6.3 to 9.8) |  |  |

IDC, invasive ductal carcinoma; ILC, invasive lobular carcinoma; MDLC, mixed ductal and lobular carcinoma; NA, not available; *P*, *P*-values of parallelism test for the within group differences (<0.05 indicates different slopes/trends compared with the reference); APC, annual percentage change; CI, confidence intervals; symbols indicate the diagnosis year corresponding to a joinpoint when two slopes/trends intercept, including § for 1998, & for 2004, ^ for 2006, \* for 2007, # for 2008, £ for 2009 and ¶ for 2010.

## Discussion

Among the 254,063 women in England with invasive breast cancer diagnosed during 1995-2011, the difference in age-standardized 5-year net survival of screened versus nonscreened cancers was significantly decreasing for the cancers diagnosed during 2007-2011. The downward-to-upward trend change in proportional incidence of nonscreened breast cancer in 2007 coincided with a steeper upward trend in age-standardized 5-year net survival of nonscreened invasive breast cancer, suggesting a possible association of the two trend changes. Similar associations were found in all strata of race, cancer stage, cancer grade and histology, except in Black patients or patients with stage I, stage III, or grade I cancer.

We here provide early evidence on the 16-year trends of proportional incidence of screened and nonscreended breast cancers among women in England. In contrast to our finding, a world-wide population study shows no decrease in incidence of advanced breast cancer following sustained implementation of breast cancer screening during 1980s-2000s, including no significant trends in Scotland.^1^ That study may be influenced by the lack of piece-wise linear modelling recommended by CDC guidelines,^23,24^ no data after 2007 and the difference between Scotland and England. It also defines advanced breast cancer mainly by cancer size, whereas we used clinical cancer staging, which is more widely used and adopted by Public Health England.^6^ Proportional incidence used here is adjusted to incidence of all breast cancers and in our view is more reliable than unadjusted incidence. We showed a downward trend in the proportional incidence of early-stage screened breast cancer since the beginning of the decrease in uptake of breast cancer screening in 2007. Thus, it is possible that the recent decrease in screening uptake is associated with decrease in proportional incidence of early-stage breast cancer and increase in that of late-stage breast cancer.

The quasi-experimental study reveals a novel association of the trend changes in proportional incidence of nonscreened breast cancer with the trend-changes in 5-year net survival of nonscreened breast cancer. Despite the increase in proportional incidence of advanced nonscreened breast cancers, our data show that a downward-to-upward trend change in proportional incidence of nonscreened breast cancer coincided with a steeper upward trend in net survival of nonscreened breast cancers after 2007. This finding is somewhat surprising, but consistent with an estimated 1-10% of overdiagnosis rate in the U.K.^4,25^ and an upward survival trend in England.^7,26^ In another word, breast cancer screening in England may not be as beneficial as reported before.^9,10,27^ More studies are needed to explain the novel association. Given the much higher rate of overdiagnosis (∼30%),^28,29^ and decreasing screening rate in the U.S.,^30,31^ it is very interesting to investigate whether the decrease in breast cancer screening is associated with an upward trend in relative/net survival in the U.S.

We also explored the factors associated with the link between increasing proportional incidence of nonscreened breast cancers and their improving age-standardized 5-year net survival. First, we show a steeper upward trend in age-standardized 5-year net survival in all strata of race, cancer stage, grade and histology among patients with nonscreened breast cancer after 2007, except in some patients. Therefore, the overall increasing survivals of breast cancer, as reported before,^7,26^ appear disproportionally linked to the nonscreended breast cancers of advanced stage, higher grade and common histologic types. Second, we found downward trends in age-standardized 5-year net survival of some screened cancers, which are grade I and other histologic types. The downward trend in these screened breast cancers is concerning and warrants more studies, but consistent with a worse overall survival of other/uncommon type of breast cancers in the U.S.^32^ Finally, patients with a black ethnical background in this study appeared not to have increasing proportional incidence of nonscreened breast cancer, nor (subsequent) steeper upward trend in net survival after 2007. However, the role of socioeconomic disparity/inequality in the screening uptake and survival of breast cancer remains controversial for English patients, including studies of supportive^33-36^ and undermined conclusions.^37,38^ More research is required to understand this role of socioeconomic disparity.

This study has several strengths. Age-standardization is critical for long-term trend analysis.^24,39^ Our findings on age-standardized net survival are consistent with the recent data of net survivals of invasive and in-situ breast cancers reported by Public Health England.^6^ Moreover, we used the most updated life tables for computing net survivals, which were levied on the recent methodological changes and advantages.^17^ Specifically, the updated life tables have better coding, enhancement to inclusion and cohort-selection criteria, and correction to capturing dates of death. In addition, subgroup analyses by race, histology, stage and grade help better understand trends among the strata of these variables. But future multivariable studies are needed to adjust for these variables if possible. Furthermore, this population-based, large scale study had sufficient statistical power and few biases, despite its limitations. Finally, the quasi-experimental design, although not as rigorous as randomized clinical trials, provides solid evidence on the association of trend changes in nonscreened breast cancer proportional-incidence with those in their net survivals.

This study has several limitations. First, survival analysis on the effects of cancer screening may result in lead-time and length-time biases. However, this quasi-experimental study was focused on the association of trend changes in the proportional incidence and net survivals of nonscreened breast cancers, and should be less susceptible to the biases. Moreover, given additional survival benefits of screened cancers linked to these biases, the decrease in net-survival benefits of screened cancers would be more profound should these biases be eliminated. Second, several prognostic factors of breast cancer and socioeconomic factors are not available for analysis including statuses of estrogen and progesterone receptors and patient income levels. Third, age was not analyzed as an exposure. Our reasoning is that, given age-standardized data, the age’s influence in the trend analysis would be minimal if any present. Fourth, due to the minimal follow-up time of 5 years for 5-year survival, we could not analyze the trends after the publication of 2012 independent review on breast cancer screening,^4^ although no immediate post-publication changes in the uptake of breast cancer screening were identified in the U.K.^40^. Finally, some cases might be misclassified histologically or clinically, although the cancer database has been widely used,^14,15,39^ and rigorously scrutinized for quality assurance.^6^

In summary, we report a downward-to-upward trend change in proportional incidence of nonscreened breast cancer among women in England in 2007. Our quasi-experimental study also shows such a trend change is associated with a steeper upward trend in age-standardized 5-year net survival of nonscreened breast cancers after 2007. The associations slightly differed by cancer characteristics and patient race. The net-survival difference of screened versus nonscreened cancers also narrowed during 1995-2011. Our findings therefore suggest decreasing survival benefits of breast cancer screening in this cohort during 2007-2016, and support reduction of breast cancer screening in some patients. Further validations are warranted.

## Data Availability

We cannot share the original data which are confidential, and were obtained with review and approval from the Public Health England.

## Authors’ contributions

HW, JB, and LZ designed the study, HW, KW and JB extracted the data, HW, SL and LZ analyzed the data, HW and LZ wrote the first draft of the manuscript, and all authors edited and approved the final manuscript.

No conflict of interest is declared by any of the authors.

We cannot share the data which are confidential and were obtained with review and approval from the Public Health England.

No funding was reported.

